# Lessons from mainland China’s epidemic experience about the growth rules of infected and recovered cases of COVID-19 worldwide

**DOI:** 10.1101/2020.04.16.20067454

**Authors:** Chuanliang Han, Yimeng Liu, Jiting Tang, Yuyao Zhu, Carlo Jaeger, Saini Yang

## Abstract

The novel coronavirus disease (COVID-19) that emerged at the end of 2019 has been controlled in mainland China so far, while it is still spreading globally. When the pandemic will end is a question of great concern. A logistic model depicting the growth rules of infected and recovered cases in mainland China may shed some light on this question. We extended this model to 31 countries outside China experiencing serious COVID-2019 outbreaks. The model well explained the data in our study (R^2^ ≥ 0.95). For infected cases, the semi-saturation period (SSP) ranges from 63 to 170 days (March 3 to June 18). The logistic growth rate of infected cases is positively correlated with that of recovered cases, and the same holds for the SSP. According to the linear connection between the growth rules for infected and recovered cases identified from the Chinese data, we predicted that the SSP of the recovered cases outside China ranges from 82 to 196 days (March 22 to July 8). More importantly, we found a strong positive correlation between the SSP of infected cases and the timing of government’s response, providing strong evidence for the effectiveness of rapid epidemic control measures in various countries.

## 1. Introduction

In December 2019, a series of cases with unknown cause of pneumonia was reported in Wuhan, the capital of Hubei Province in mainland China (WHO_a_). Later, deep-sequencing analysis of lower respiratory samples confirmed the presence of a novel coronavirus, which was firstly named as 2019 novel coronavirus (2019-nCoV) on Jan 12, 2020 and may have originated from certain bats (Zhou et al., 2020). The coronavirus (SARS-CoV-2) was officially renamed as COVID-19 on February 12, 2020. Human-to-human transmission through of COVID-19 has been confirmed not only in China (Wang et al., 2020; Li et al., 2020; Zhu et al., 2020; Hui et al., 2020), but in countries around the world such as the Republic of Korea (Choi et al., 2020; Ki et al., 2020; Shim et al., 2020), Italy (Livingston et al., 2020; Spina et al., 2020; Rosenbaum, 2020) and Iran (Tuite et al., 2020; Abdi, 2020; Zhuang et al., 2020). At the time of writing, the epidemic situation in mainland China has been effectively controlled since there are only sparse daily local new cases since Mar 19, 2020, and the recovery rate in mainland China has risen up to 93.51% by April 13 (Chinese Center for Disease Control and Prevention, CCDC). However, the epidemic situation outside China is worsening. As of April 13, 2020, globally, a total of 1788665 confirmed cases of COVID-19, and death and recovered rates of 6.35% and 20.45 %, respectively, have been reported (Sina.com). When the inflection point will appear for both infection and cure outside China remains unclear. The hope and lessons on COVID-19 control from China is necessary and worth quantifying (Azman & Luquero, 2020).

We applied a descriptive model that has been proven to be robust and stable (Han et al., 2020) to analyze the global data on COVID-19 cases globally (infected and recovered cases in mainland China, and infected cases countries outside China). We then estimated the relation between parameters of infected and recovered cases in mainland China. Next, we used that relationship to map the parameter space of infected case to the parameter space of recovered cases for 31 countries globally. Finally, we explored the relationship between the model parameters and governmental control measures.

## 2. Method

### 2.1 Sources of Data

The cumulative number of confirmed and recovered COVID-19 cases in mainland China was obtained from the National Health Commission of China, and the Provincial Health Commissions of 30 provincial administrative regions (excluding Tibet, because the only one confirmed infected COVID-19 case in Tibet has recovered on Feb. 12) in mainland China (January 10 to March 19, 2020). The data are publically available. All cases were laboratory confirmed following the published standards made by the National Health Commission of China (CCDC_a_). The basic test procedure has been described in detail in previous work (Zhou et al., 2020; Huang et al., 2020). This dataset was partially analyzed in an initial work on this topic (Han et al., 2020).

For our analyses we selected 31 countries outside China with serious coronavirus epidemic situations (January 10 to April 13, 2020). Their population amounts to 39.4% of the world’s population (57.8% with China included) and 81.6% of the world’s infected COVID-19 cases (86.2% with China included) (WHO_a_). The data of COVID-19 cases in countries outside China were obtained from the situation reports on the official website of the World Health Organization (WHO_a_), which is publicly available. The data used in this study include the cumulative number of reported laboratory-confirmed COVID-19 cases. Countries included in our study are: Republic of Korea, Japan, Australia, and Singapore, all in the Western Pacific Region; Italy, Spain, France, Germany, Switzerland, the United Kingdom, Netherlands, Sweden, Denmark, Austria, Belgium, Portugal, Czech Republic, Finland, Ukraine, Slovakia, Bulgaria, Lithuania, in Europe; India in South-East Asia; the Islamic Republic of Iran, Lebanon, in the Eastern Mediterranean Region; the United States of America, Canada, Brazil, Mexico, in the Americas; South Africa, Ethiopia, in Africa. All the laboratory-confirmed cases are determined according to the WHO standard. The overnments of these countries all declared a state of national emergency, of wartime, or a blockade of their borders as a result of the COVID-19 outbreak. In this study, we collected the beginning of government intervention from the mainstream authoritative media of each country selected.

### 2.2 Epidemic curve modeling

In this work, we used a sigmoid descriptive model (Equation (1)), as derived from logistic differential equations (Han et al., 2020). 

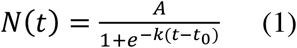

*N*(*t*) is the general form of the cumulative numbers of infected or recovered patients at time *t. A* denotes the maximum number of infections or recoveries, *k* is the logistic growth rate, *t*_*0*_ is the inflection point of the sigmoid curve. This descriptive model has been verified by not only with the infected, death and recovered cases of COVID-19, but also with the data of SARS in 2003 (Han et al., 2020). For the infected cases, there are three parameters (*A, k, t*_*0*_) in the model; for the recovered cases, we fixed *A* to the difference between the maximum number of cumulative infections and deaths, based on biological fact. It is worth mentioning that in our model, *t*_*0*_ is the mathematically defined inflection point. In this paper, we assumed that *t*_*0*_ is the time of inflection of the epidemic dynamics in a region.

We processed the data and modeled them with custom scripts on MATLAB (the Math Works). We adopted the nonlinear least square (NLS) algorithm for data fitting and parameter estimation.

## 3. Results

Based on the cumulative number of confirmed and recovered cases in 30 provinces of mainland China as well as in 31 countries, with the above-mentioned model all data could be well explained (*R*^2^>0.95) (See Methods, Fig. 1A-C, Fig. 4A-C).

**Figure 1.**
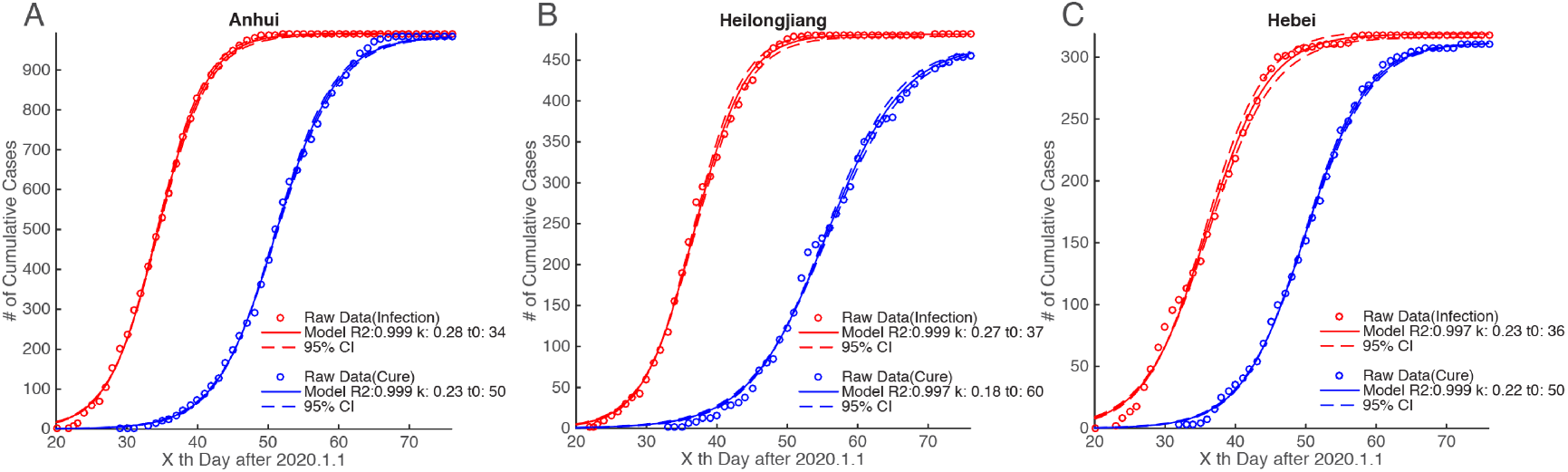
Example of provinces for the time series of infected and recovered cases with corresponding fitted curve in mainland China A-C show cases in three provinces (Anhui, Heilongjiang and Hebei). The horizontal axis in each panel denotes the x^th^ day after January 1, 2020. The vertical axis in each panel is the cumulative number of infected (red circles) and recovered cases (blue circles). The red and blue line are the fitting curves of the sigmoid model, and the dashed red and blue curves mark the 95% confidence interval of the fitting curves.

**Figure 2.**
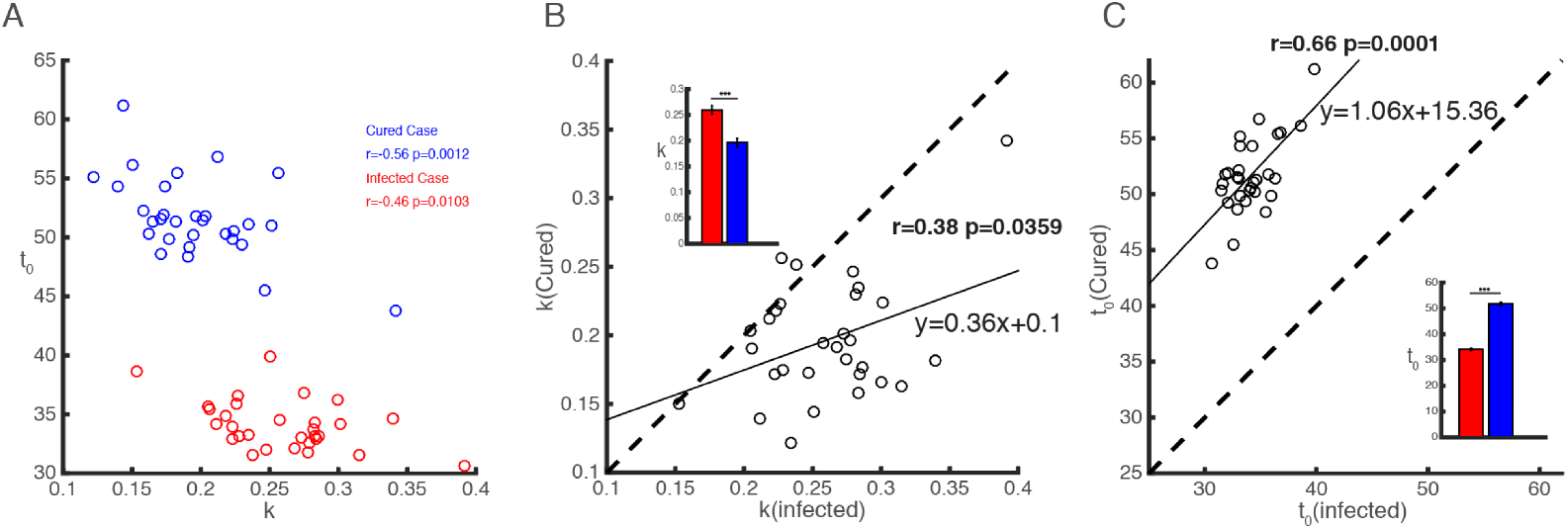
Relationship between the rules for infected and recovered cases in mainland China Panel A shows the scatter plot of the logistic growth rate (k) and the semi-saturation period (t_0_) for infected (red circles) and recovered cases (blue circles). Panel B is the scatter plot of the logistic growth rate (k) of infected and recovered cases (black circles), the dashed line is the diagonal line of the coordinate system (the same for Panel C). The relation between the logistic growth rates of infected and recovered cases is modeled with a linear function, which is shown as the black line. The bar graph in the northwest corner shows the difference between *k* for infected and for recovered cases. Panel C shows the scatter plot of the semi-saturation periods (t_0_). The black curve is the linear function estimated based on the relation between infected and recovered recovereed. The bar graph in the southeast corner shows the difference between t_0_ for infected and recovered cases.

**Figure 3.**
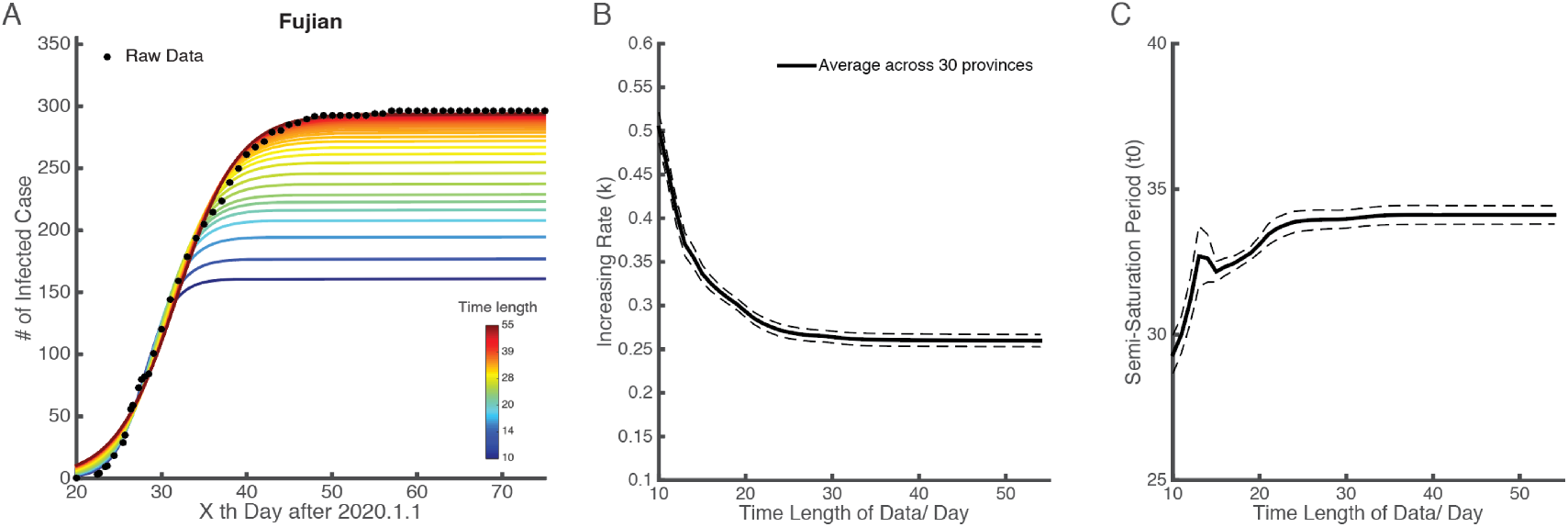
Time stability of the logistic model for infected cases in mainland China Panel A shows the example of Fujian province. As in panels B and C, the horizontal axis denotes the × th day after January 1, 2020. The vertical axis in panel A indicates the cumulative number of infected (black dots). The lines with different colors are the fitting curve by the logistic model using data with different time length as shown in color bar in the southeast corner. Panels B and C illustrate how the estimated parameters (k and t_0_) change with increase time length of the availble data. In panel B, the black curve shows the average logistic growth rate (k) across 30 provinces in mainland China, depending on the time length of data. The dashed lines mark the standard error. Panel C does the same for the semi-saturation period

**Figure 4.**
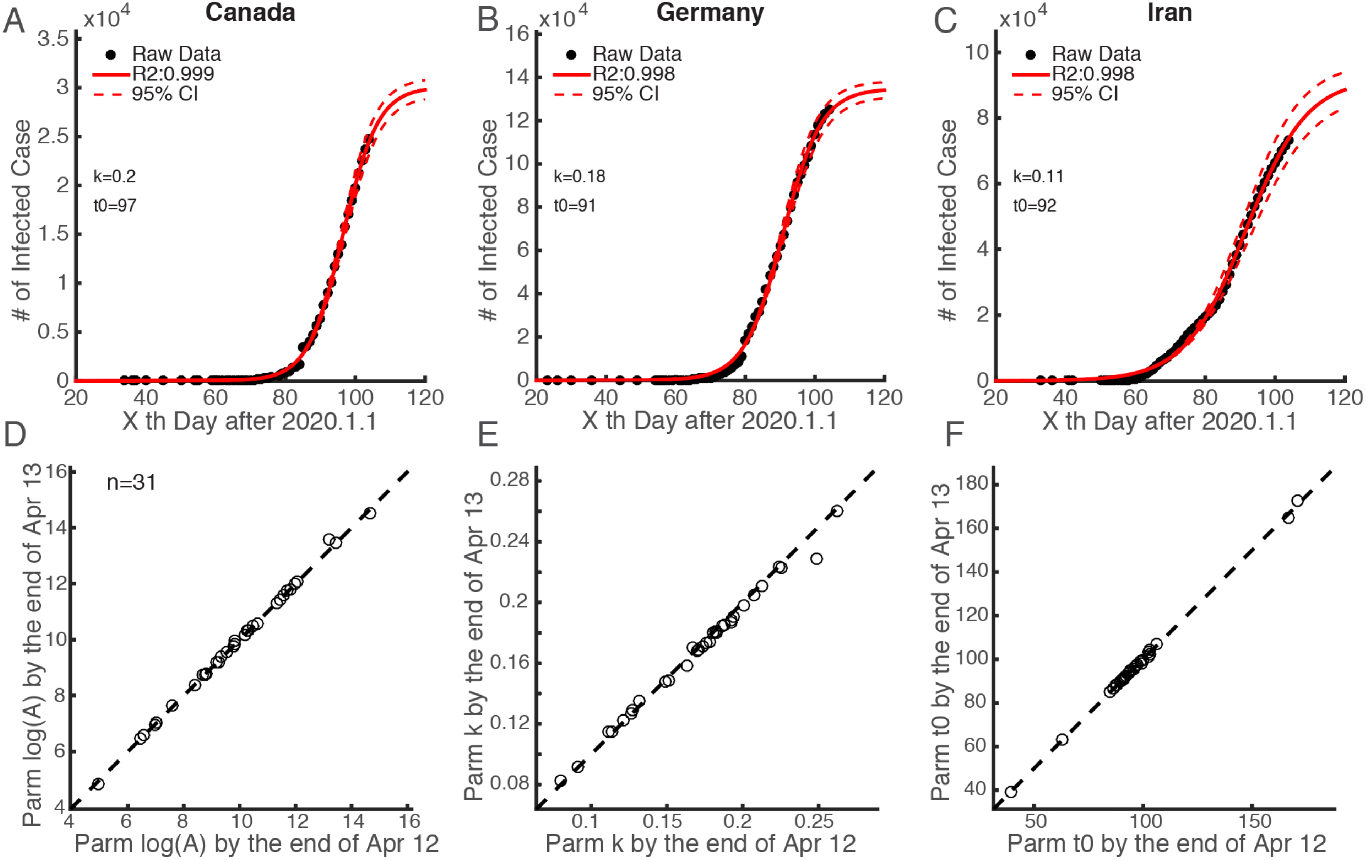
Example countries for the time series of infected cases with corresponding fitted curves around the world and their temporal stability Panel A-C show infected cases and their fitted curves for Canada, Germany and Iran. The horizontal axis is the x^th^ day after Jan 1, 2020. The vertical axis indicates the number of infected cases in the corresponding country. The black dots in each panel are raw data of the cumulative number of infected cases. The red line is the fitted curves by our descriptive model. The dashed red lines mark the 95% confidence interval of the fitted curves. Panel D-F show the difference of parameters estimated from relative longer (by the end of Jun 13) and shorter (by the end of Apr 12) time series data, each dot denotes for one country.

### 3.1 Relationship between infected and recovered cases in mainland China

The epidemic situation of COVID-19 has been controlled in China (CCDC_b_), with a recovery rate of 93.51% (CCDC_a_). The time series of infected and recovered cases and their fitted curves for the three example provinces (Anhui, Heilongjiang, Hebei) are shown in Fig. 1A-C. By observing the fitted curves of all provinces in mainland China (Supplementary Figure S1), we found that the logistic growth rate (*k*) of infected cases is larger than that of recovered cases, and that the semi-saturation period (*t*_*0*_) of infected cases is shorter than that of recovered cases.

As shown in Figure 2, the parameter spaces of *k* and *t*_*0*_ are different for infected (red circle) and recovered cases (blue circle), and there is a significant negative correlation between the two (for infected cases, *r*=-0.46, *p*=0.0103; for recovered cases, *r*=-0.56, *p*=0.0012; Pearson correlation). When we compared these two parameters of infected and recovered cases, we found that infection cases have larger (*t*=6.9136, *p*<10^−7^, right-tailed test) growth rates (Fig. 2B) but smaller (*t*=-37.271, *p*<10^−25^, left-tailed test) semi-saturation periods (Fig. 2C) than recovered cases. The growth rates of infected and recovered cases is significantly positively correlated (*r*=0.38, *p*=0.0359, Pearson Correlation), and so is the semi-saturation period of infected and recovered cases (*r*=0.66, *p*=0.0001, Pearson Correlation). Each of them could be fitted to a linear model, with slope and intercept of 0.36 (CI: [0.0268, 0.698]) and 0.1 (CI: [0.014, 0.191]), respectively (panel B), while those of panel C are 1.06 (CI: [0.59, 1.54]) and 15.36 (CI: [-0.85, 31.56]).

### 3.2 Growth rules of COVID-19 infections outside China

The dataset of infected case in mainland China shows the time stability of the logistic model (Fig. 3). The example of the province of Fujian is shown in Figure 3A. It is clear that as the in length of the time-series increases, the fitting curves converge to the actual number of cases (black dots).

We defined an index for minimum time length (MTL) to measure the amount of data necessary to produce a stable output. This index is the time length at which the change of parameter value (*k* or *t*_*0*_) is less than 5% in two consecutive days. With the data of 30 provinces in China, we found that the MTL for the logistic growth rate (*k*) is 15 days (Fig. 3B) and for the semi-saturation period (*t*_*0*_) 26 days (Fig. 3C). This result indicates the amount of data required to measure the epidemic situation.

After identifying the relationship between the increasing rate and the semi-saturation period, we applied this relationship to the COVID-19 cases from 31 countries. We applied the descriptive model to fit the data for cumulative number of infected cases in each of the 31 countries. Figure 4 shows the time-series data of infection for three countries (Canada, Germany and Iran, Fig. 4A-C; see the supplementary Figure S2 for all 31 countries). All data series for infected cases could be well explained by our model (*R*^2^>0.95). The average change rates of the parameters are already lower than 5% (Fig 4 D-F), which indicates that the parameters we estimated from the model are stable. We noticed that the logistic growth rate of 31-country cases does not have a significant difference (*t*=1.197, *p*=0.237 two-tailed test) with that of Chinese provinces, but the 31-country cases have a significantly longer (*t*=-17.4819, *p*<10^−20^, right-tailed test) semi-saturation period. The numbers indicate that the t_0_ of infections in 31 countries has an average of 98 days (std: 20.5), and the average date matched to this is around April 7 (ranging from Mar 3 to Jun 18).

### 3.3 Predictions for the growth rules of recovered COVID-19 cases and their relation to the timing of governmental control measures

On observing the logistic growth rate (*k*) and the semi-saturation period (*t*_*0*_) of infected cases in 31 countries, we found there was a negative correlation (r=-0.63, p=0.0001, Pearson Correlation) (Fig. 5) between these two parameters, which shows a similar tendency to the results of provinces in mainland China (Fig. 2A). To predict the logistic growth rate (*k*) and the semi-saturation (i.e. inflection) point (*t*_*0*_) of the recovered COVID-19 cases outside China, we mapped the parameter space of infected cases into the parameter space of recovered cases, based on the parameter relationship between infected cases and recovered cases obtained from China (Fig.2BC). For the 31 countries, the logistic growth rate ranges from 0.08 to 0.45. The mean *t*_*0*_ of the fitted curves for recovered cases is 119.71 (std: 21.73), which means that the mean semi-saturation period (*t*_*0*_) for these 31 countries will arrive on, approximately April 29, 2020 (ranging from Mar 22 to July 8).

**Figure 5.**
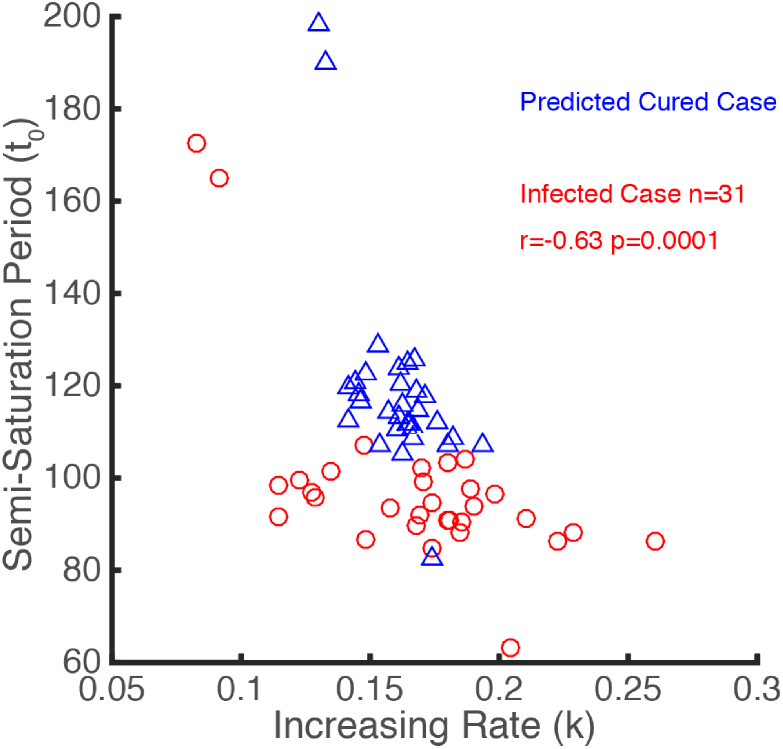
Prediction for growth rules of recovered cases around the world based on the Chinese experience The scatter plot of the logistic growth rates (k) and the semi-saturation periods (t_0_) for infected cases is showm in red circles. Based on the linear model shown in Fig 2BC, predictions of k and t_0_ for recovered cases are plotted as blue triangles.

To explore the practical implication of these parameters, we took the timing of governmental emergency control measures related to COVID-19 as a variable to analyze its relationship with the key parameters (Fig. 6). We found that the semi-saturation time showed a significant positive correlation (*r*=0.73, *p*<0.0001, Spearman correlation) with the timing of governmental control measures (Fig. 6 BE). k showed a marginally significant negative correlation (*r*=-0.3, *p*=0.0925, Spearman correlation), while the logistic growth rate or the maximum of infected casesa do not show a strong correlation (*r*=0.02, *p*=0.91, Spearman correlation) with the timing of control measures (Fig. 6 ACDF). China released the earliest national control measures to prevent the spread of COVID-19 (on Jan. 23) and its *t*_*0*_ is the lowest (Figure 5, red circle). We also noticed that the timing of governmental emergency control measures is significantly earlier than the semi-saturation period (Fig. 6B) (*t*=2.99, *p*=0.0043, two-tailed test), which indicates that most of the selected countries have taken measures in the early phase of the outbreak.

**Figure 6.**
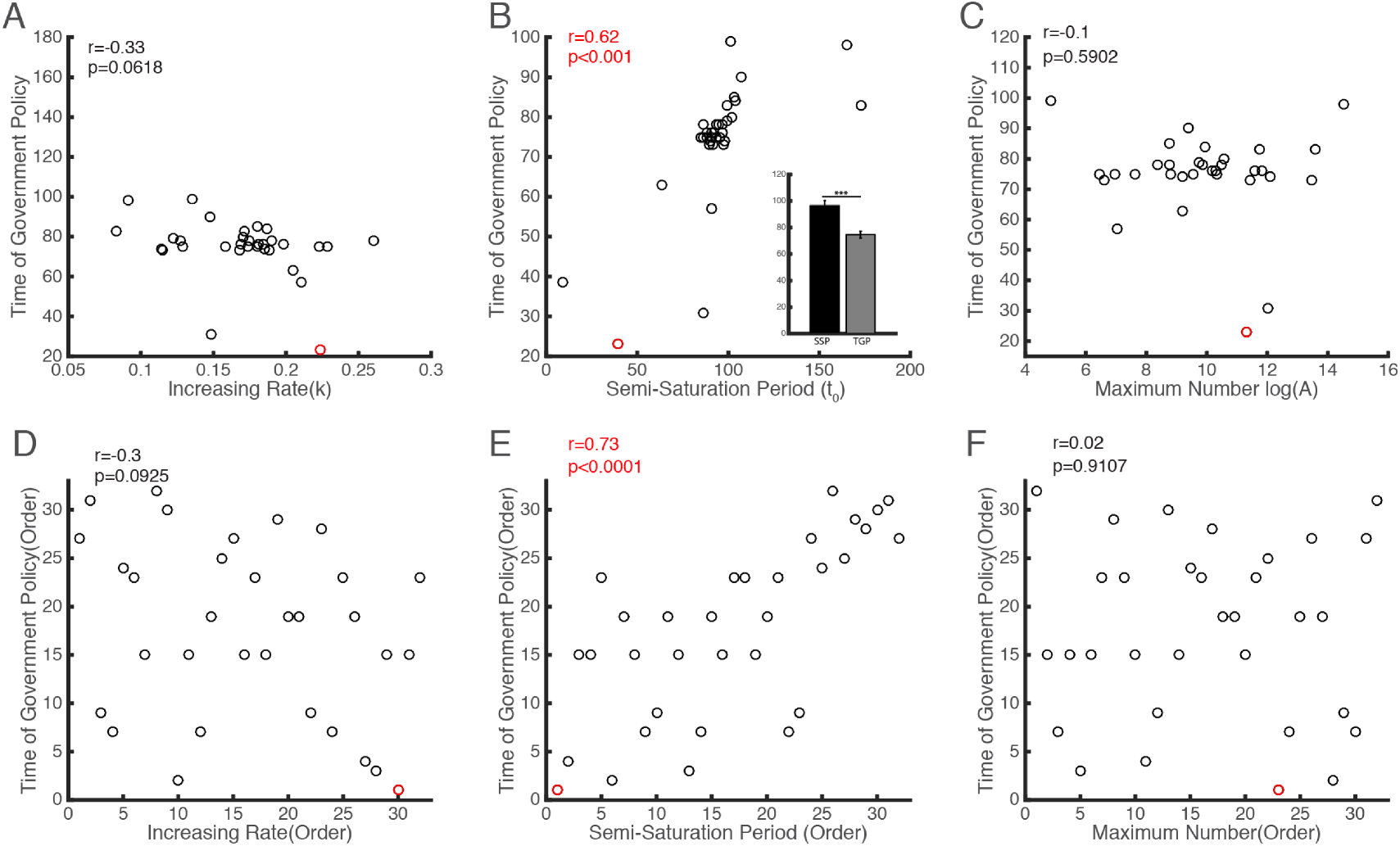
Relationship between the timing of government intervention and the model parameters of infected case for each country Panel A-C show the scatter plot of the time of government measures and the model parameters of infected case (k, t_0_ and A, respectively). In the southeast corner of Panel B, a bar graph was shown to indicate the difference between timing of government measures (grey bar) and semi-saturation period (SAP, black bar). D-F shows the same data of A-C but in the order form. The red circle in six panels denotes the model parameters in China’s infected case.

## 4. Discussion

Based on data for 30 provinces in mainland China from Jan 20 to Mar 29, 2020, we have showed that the the growth characteristics of infected and recovered cases, are positively correlated (Fig. 2 BC). Further, we explained the data from 31 countries using the same model (see methods). We predicted the logistic growth rates (*k*) and semi-saturation times (*t*_*0*_) of recovered cases based on the knowledge of data from China. It is worth mentioning that there is a strong correlation between these parameters and the timing of governmental emergency policies on COVID-19.

### 4.1 Comparison with previous work

Different from models that have been applied to COVID-19, like SIR or SEIR (Li et al., 2020; Wang et al., 2020; Tian et al., 2020), our model is relatively simple, but robust. The present investigation is an extension of our previous work (Han et al., 2020). We have verified that our model explains the data in various geographic spaces of mainland China well, and we have provided robust results with different lengths of time series.

This model is able to capture the macroscopic dynamics of the epidemics worldwide (*R*^2^>0.95) (Fig. 4), and we quantitatively investigated the minimum data length to produce a stable output (Fig. 3). By now, the number of domestic infected cases in China is very limited, which is consistent with our prediction results. The cumulative numbers of infected and recovered cases are almost saturated. This means that the spread of the virus has been effectively controlled.Thus we believe that the parameters estimated in this work for Chinese provinces are reliable (Fig. 1A-C). The characteristics of two curves can depict the dynamic macroscopic features of the epidemic situation.

Moreover, we extended this model to the data in each of 31 countries (Fig 4 A-C), and found that the sigmoid model fits the data very well, too (*R*^2^>0.95) (Fig 4 A-C).

### 4.2 Relationship among model parameters and the prediction of the recovered cases

We noticed that the semi-saturation period of infected and recovered cases in China showed a positive correlation (*p*-value = 0.0001). This phenomenon corresponds to common sense, since recovery is always later than infection. Although the positive correlation between the logistic growth rates of infected and recovered cases is significant (*p*-value = 0.0359), the correlation is much lower than that of semi-saturation times. This result is reasonable, too, since the causes determining the logistic growth rates of infected cases are quite different from those of recovered cases.

When comparing *k* and *t*_*0*_ in specific cases, no matter whether in mainland China or countries worldwide (Fig. 5), we found significant negative correlations. This is consistent with our initial work (Han et al., 2020). As for the negative correlation between *k* and *t*_*0*_ for recoveredcases, it might also be as the casethat when the logistic growth rate (*k*) of the recovered cases is larger, it will take less time to curemost patients.

### 4.3 Suggestion for governments around the world on the prevention and control of COVID-19

Most importantly, we found there is a strong correlation between the timing of governmental control measures for COVID-19 in 31 countries and the semi-saturation period estimated (Fig 6 BE). This indicates that the early implementation of the government’s prevention and control policy effectively shortened the turning point of the epidemic. Taking China as an example, with regard to the COVID-19 outbreak in Wuhan the strict isolation policy was announced on Jan. 23, and the inflection point there was around Feb 9, 2020, so that the time lag was 17 days. Across all countries, the timing of governmental policy is significantly shorter than the semi-saturation period of infected cases, which is also a strong signal for governments of all countries to take control measures as early as possible.

## Data Availability

All data is publically available on the WHO website

https://www.who.int/docs/default-source/coronaviruse/situation-reports

## Acknowledgements

This study was sponsored by the National Key Research and Development Program of China (2018YFC1508903), the National Natural Science Foundation of China (41621061) and the support of International Center for Collaborative Research on Disaster Risk Reduction (ICCR-DRR).

---

**Supplementary Figure S1.**
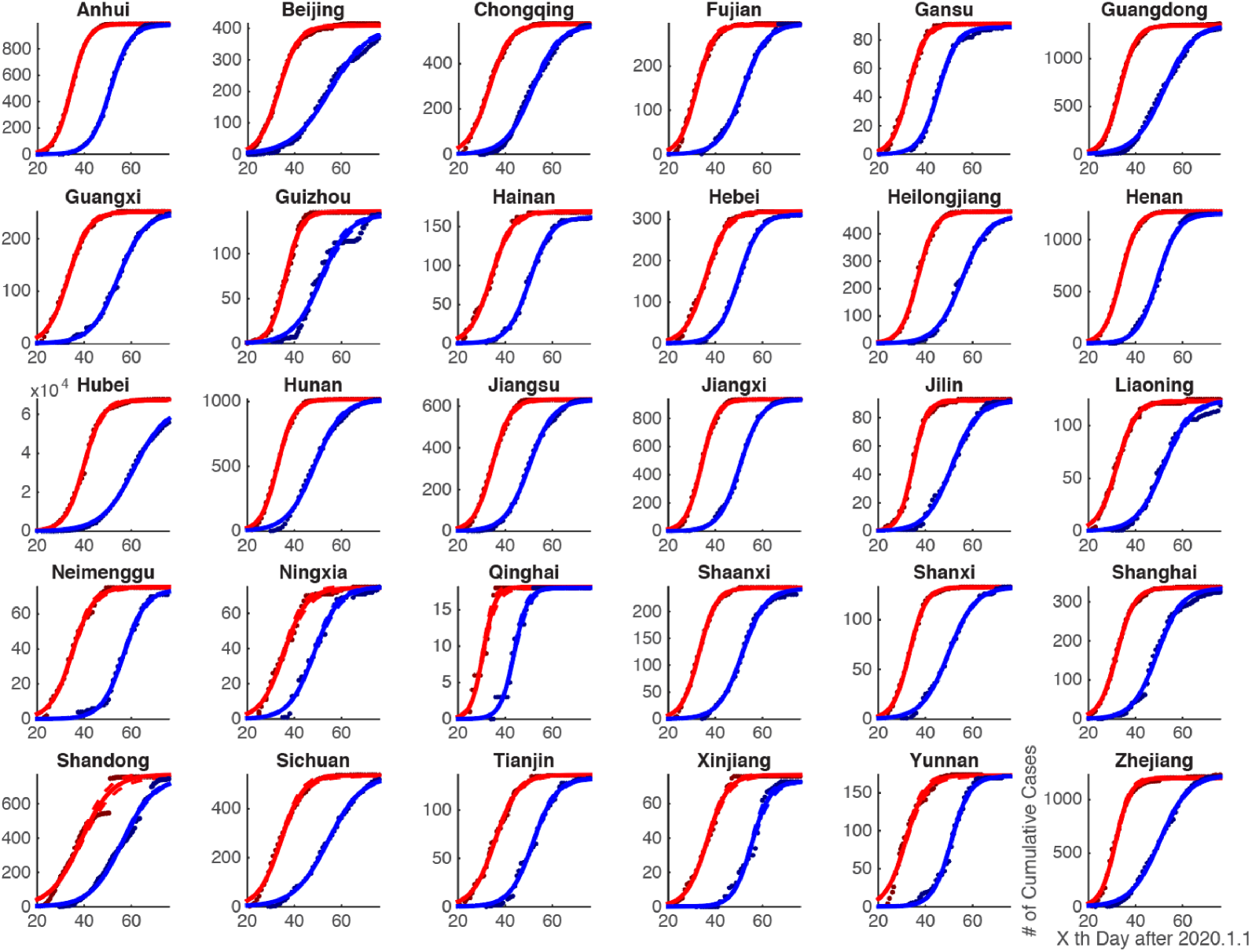
Intrinsic growth rules of patients infected with 2019 novel coronavirus in all provinces of mainland China. The deep red and blue dots in each panel are raw data of the cumulative infected and recovered cases. The red and blue lines represent the curves fitted by our descriptive model.

**Supplementary Figure S2.**
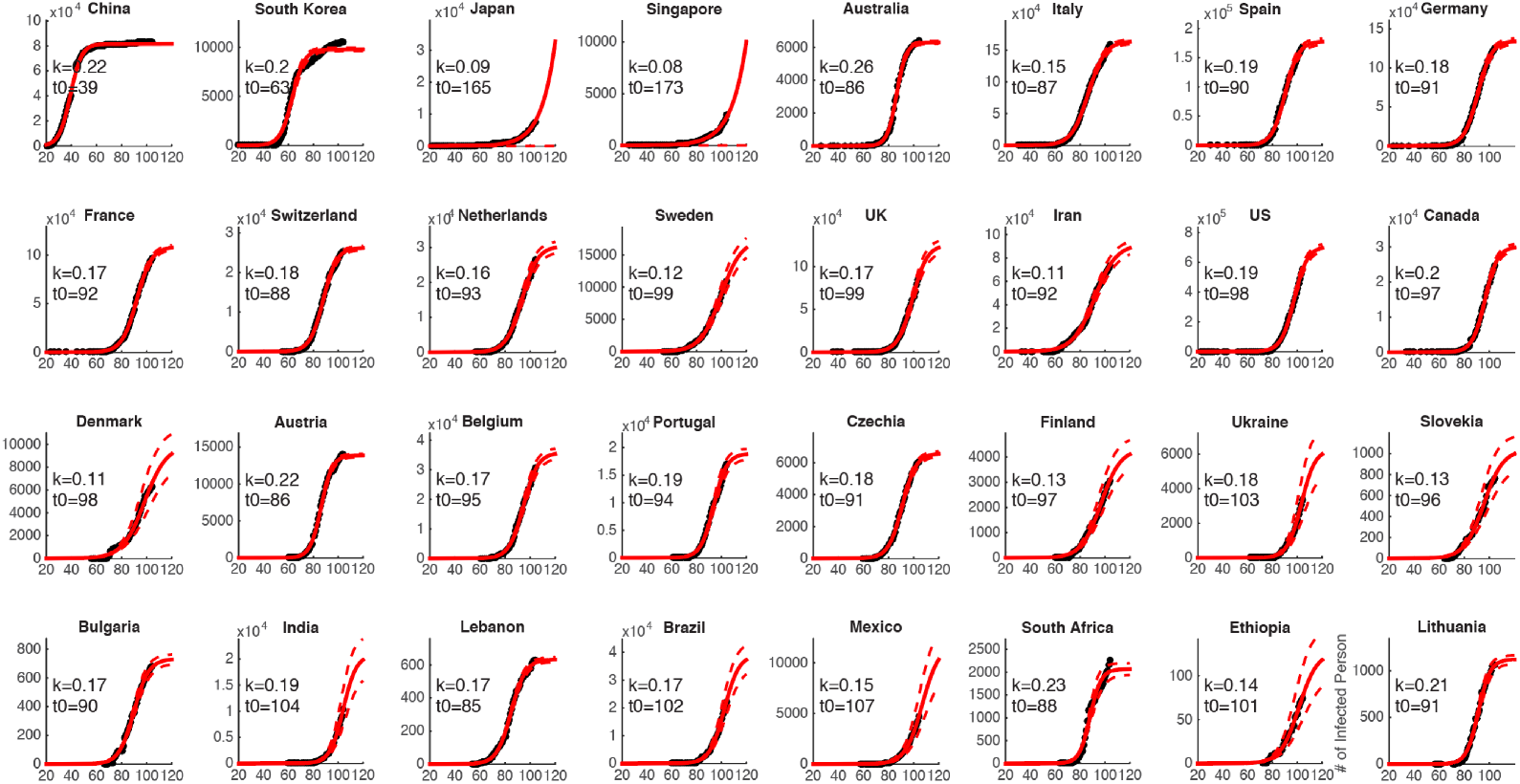
Intrinsic growth rules of patients infected with 2019 novel coronavirus in 31 countries around the world. The black dots in each panel represent the raw data of the cumulative infected cases. The red line is the curve fitted by our descriptive model.

